# Predominance of Distinct Autoantibodies in Response to SARS-CoV-2 Infection

**DOI:** 10.1101/2021.09.14.21263603

**Authors:** Yunxian Liu, Joseph E. Ebinger, Rowann Mostafa, Petra Budde, Jana Gajewski, Brian Walker, Sandy Joung, Manuel Bräutigam, Franziska Hesping, Elena Schäfer, Ann-Sophie Schubert, Hans-Dieter Zucht, Gil Y. Melmed, Kimia Sobhani, Jonathan Braun, Dermot P.B. McGovern, Jennifer E. Van Eyk, Susan Cheng, Justyna Fert-Bober

## Abstract

**Background:** Improved knowledge regarding the prevalence and clinical significance of the broad spectrum of autoantibodies triggered by SARS-CoV2 infection can clarify the underlying pathobiology, enhance approaches to evaluating heterogeneity of COVID-19 clinical manifestations, and potentially guide options for targeting immunosuppressive therapy as the need for more effective interventions continues to evolve. In this study, we sought to determine the prevalence of autoimmune antibodies in diverse cohort of SARS-CoV-2 positive healthcare workers and measure the extent to which factors associated with triggered autoimmunity are activated even following mild and asymptomatic infection.

**Methods:** Antigen microarrays were used to profile reactivity of IgG autoantibodies against 91 proteins and cytokines based on autoantibody profiling studies in autoimmune diseases.

**Results:** In this discovery screening study, we found that 90% of the IgG positive individuals demonstrated reactivity to at least one autoantibody. When compared to results of the same assays conducted on samples from pre-COVID-19 controls, our primary cohort of individuals with SARS-CoV-2 IgG antibody positivity had significantly elevated IgG against twelve additional proteins including CHD3, CTLA4, HARS, IFNA4, INS, MIF, MX1, RNF41, S100A9, SRP19, TROVE2, and VEGFA. These findings confirmed that all severity levels of SARS-CoV-2 infection, even asymptomatic infections, trigger a robust and diverse autoimmune response; our results also highlight the utility of multiparametric autoantibody detection in this setting.

**Interpretation:** Taken together, our findings underscore the serological diversity underlying the clinical heterogeneity of COVID-19 infection and its sequelae, including the long-Covid phenotypes.

**Funding:** This work was supported in part by Cedars-Sinai Medical Center (JEE; SC), the Erika J Glazer Family Foundation (JEE; JEVE; SC), CSMC Precision Health Grant (JFB), the F. Widjaja Family Foundation (JGB, GYM, DM), the Helmsley Charitable Trust (JGB, GYM, DM), and NIH grants K23-HL153888 (JEE) and DK062413 (DPBM).

**RESEARCH IN CONTEXT:** *Evidence before this study:* Currently, several studies have shown the possible involvement of autoimmunity in patients affected by coronavirus disease 2019 (COVID-19). In contrast to cytokine storms, which tend to cause systemic, short-duration problems, autoantibodies (AABs) are thought to result in targeted, longer-term damage and development of autoimmune diseases.

*Added value of this study:* According to our knowledge, we evaluated the largest number of protein antigens to characterize the prevalence and heterogeneity of the AABs signature in SARS-CoV-2 convalescent individuals. We examined autoimmune reactivity to SARS-CoV-2 in the absence of extreme clinical disease to acknowledge the existence of AABs even among those who had mild-to-moderate or no symptoms during their illness, as a hallmark of ongoing long-COVID syndrome. Through our analysis we suggest that VEGFA, MIF, IFNA4, SPP1 and APOH could be used as hallmark for SARS-CoV-2 infection and activation of the autoimmune system.

*Implications of all the available evidence:* Our study comprehensively characterized the heterogeneity of the AABs signature in SARS-CoV-2 convalescent individuals. The results established a list of diagnostic signatures and potential therapeutic targets for long-Covid-19 patients although follow-up long-term studies are required. We believe that our findings will serve as a valuable resource, to drive further exploration of long-COVID syndrome pathogenesis.

## INTRODUCTION

COVID-19 manifests with a broad spectrum of clinical phenotypes that are characterized by accentuated and misdirected host immune responses (1-3). While pathological innate immune activation is well documented during various stages of COVID-19 (4), the impact of autoantibodies (AABs) on disease progression is less defined. Recent reports have detected reactivity of several AABs among subsets of more severely affected COVID-19 patients, including those that are characteristic of classic systemic autoimmune diseases, such as systemic lupus erythematosus (5), Guillain Barre syndrome, (6, 7), vasculitis (8), and multiple sclerosis (9). Although some disease-modifying AABs responses have been described in patients with critical COVID-19, for example AABs against type I IFNs (10) or anti-phospholipid antibodies (1), the full breadth of AABs reactivities in COVID-19 and their immunological and clinical importance remain unclear.

The aim of this study is to comprehensively characterize the diversity of SARS-CoV-2 AAB production against auto-antigens that have been previously linked to classic autoimmune diseases. Identifying and quantifying production of specific AAB to distinct antigens, representing a wide range of tissue or cellular organelles, can help to elucidate the heterogeneity of autoimmunity responses triggered by SARS-CoV-2 infection as well as their role in the onset and persistence of post-acute clinical sequelae of COVID-19.

## METHODS

### Study design and participants

The sampling strategy for our study has been described previously (11). In brief, beginning on 11 May 2020, we enrolled a total of n=6318 active employees working at multiple sites comprising the Cedars-Sinai Health System, located in the diverse metropolis of Los Angeles County, California (healthcare workers, HCWs). For all HCW participants, EDTA plasma specimens were transported within 1 hour of phlebotomy to the Cedars-Sinai Department of Pathology and Laboratory Medicine and underwent serology testing using the Abbott Diagnostics SARS-CoV-2 IgG chemiluminescent microparticle immunoassay (Abbott Diagnostics, Abbott Park, Illinois) against the nucleocapsid (N) antigen of the SARS-CoV-2 virus (12, 13). For the current study, we included n=177 participants who were IgG positive and had completed electronic survey forms via Research Electronic Data capture (REDCap) (14). Survey forms collected data on pre-existing traits, self-reported symptoms, and medical history characteristics (**Supplemental Table 1**). Data on the following systemic symptoms were collected and recorded: chest pain, chills, conjunctivitis, dry cough, productive cough, diarrhea or other digestive symptoms, fatigue, fever, headache, loss of appetite, muscle aches, nasal congestion, nausea, runny nose, shortness of breath, skin changes, loss of smell/taste, sneezing, sore throat, vomiting, and reports of all other symptoms. For each symptom, we collected data at the time of the blood draw on the timing of any concurrent or previously experienced symptoms; these data were categorized as follows: 0 = never symptoms, 1 = within the last 6 months, 2 = within the last 3 months, 3 = within the last 2 months, 4 = within the last 1 month, 5 = currently or within the last week. The relative timing, in relation to the blood draw, was scored for each symptom and then the sum of scores for all reported symptoms was calculated as the total symptom burden for each HCW. The HCWs with a total symptom burden score lower than or equal to 22 were then categorized into the ‘earlier’ group, and HCWs with a score of 22 or higher were grouped into the ‘recent’ group. All study participants provided written informed consent and all study protocols were approved by the Cedars-Sinai Medical Center institutional review board. Age and sex matched pre-pandemic serum samples obtained from the Bavarian Red Cross (Wiesentheid, Germany) with ethical approval from the Bayerische Landesaerztekammer (Study No. 01/09) served as a healthy comparator group, i.e., healthy controls (HCs). Additionally, to serve as a positive comparator group, we obtained serum samples (n=6) from patients with systemic lupus patients (SLE) who fulfilled at least 4 of the 11 American College of Rheumatology revised criteria for the classification of SLE (West Sussex, United Kingdom).

### Luminex autoantibody array and autoantigens design

A total of 91 protein antigens were selected for this study, with specific antigens prioritized based on their previously reported associations with SLE (15) and viral infection (16). The panel includes cytokines, lung-specific proteins, protein partners that are known to interact with COVID-19 proteins (17, 18) proteins with high amino acid homologies to COVID-19 proteins (19), as well as known auto-antigens implicated in systemic sclerosis (SSc), rheumatoid arthritis (RA), Sjögren’s syndrome (SjS), mixed connective tissue disease (MCTD) and idiopathic inflammatory myopathies (IIM) (**Supplemental Table 2**). The following antigens were purchased from Diarect AG (Freiburg, Germany): U1-snRNP68/70□kDa (SNRNP70), U1-snRNP A (SNRPA), U1-snRNP C (SNRPC), U1-RNP B/B□(SNRPB), SmD3 (SNRPD3), ribosomal phosphoprotein P0 (RPLP0), ribosomal phosphoprotein P1 (RPLP1), DNA topoisomerase I (TOP1, Scl-70), SSA/Ro52 (TRIM21), myeloperoxidase (MPO), SSB/La, and PDC-E2 (DLAT). Other antigens were produced in-house using *E*.*coli* SCS1 carrying plasmid pSE111, which contains an N-terminally located hexa-histidine-tag. A subset was expressed with a BirA and hexa-histidine-tag in *E*.*coli* BL21. Antigens were affinity-purified under denaturing conditions using Protino® Ni-IDA 1000 funnel columns (Macherey-Nagel, Düren, Germany) (15). The selected analytical targets can be categorized by functional protein families or their previously reported associations with immune-relevant clinical disease states (20) (**Supplemental Table 2**).

The SeroTag^®^ AABs workflow was designed to profile the AABs reactivity using Luminex color-coded beads technology as outlined previously (15). Briefly, for each coupling reaction up to 97□µg antigen and 4□×□105□MagPlex^™^ beads per color were used. Coupling efficiency was confirmed by incubation of 625 beads from each coupled region with a phycoerythrin-conjugated anti-6×□HisTag antibody (Abcam, Cambridge, UK) at a concentration of 10 μg/mL for 45 min shaking at 900 rpm and room temperature. Coupled beads were mixed to a final concentration of 62.5 beads/μL and stored in PBS supplemented with 1% bovine serum albumin, 0.1 % Tween 20 and 0.05% ProClin^™^ 300 (Merck KGaA, Darmstadt, Germany), at 4 °C until use. Plasma or serum samples from all subjects were diluted 1:100 in assay buffer (50% PBS, 1% BSA, 50% Low-Cross Buffer (Candor Biosciences, Wangen, Germany). Diluted samples (50ul) and beads (50ul) were mixed and incubated for 22 hrs at 4-8 °C in the dark. The bound AABs were detected by addition of R-phycoerythrin-labelled goat anti-human IgG detection antibody (Ab) (5□µg/ml, Dianova, Hamburg, Germany) for 1 hr at RT, after several PBS washes. The Luminex FlexMAP3D analyzer identified and quantified each antigen-AABs reaction based on bead color, median fluorescence intensity (MFI) and antigens fulfilling the minimum bead count criterion (>10 beads measured per bead ID). The background reactivity of the screen was determined using all healthy control samples. The 0.1 quantile of MFI values of all samples was calculated (<500 MFI). Median intra and inter-plate coefficient of variation (CV) was calculated by measuring reference samples in triplicate on all and each assay plates. The cut off applied for the median intra- and inter-plate CV was <30%. Matched analysis of samples from other convalescent subjects showed no differences in AAB values. The dynamic range was determined using blank samples and a control bead coupled to BSA (control_BSA) for the lower and huIgG (control_huIgGhi) for the upper MFI range.

### Statistical analyses

Data processing and analysis were performed using R v3.5.1 and KNIME 2.12 (https://www.knime.org/) (21, 22). Among the pre-pandemic non-SLE control group (HCs), a mean and standard deviation (STD) was calculated for the MFI for each of the 91 antibodies assayed. An individual was classified as having an increased AAB level if the MFI for that given AAB was at least 3 SD above the mean level in the pre-pandemic HC group. Frequencies of AABs and categorical variables among the groups were compared using Chi-square test. Kruskal-Wallis Rank Sum Test was conducted to compare nonnormal continuous variables (variables were summarized in median and interquartile range (IQR). Statistical significance was defined as a two-tailed P value less than 0.05.

## RESULTS

### Cohort characteristics and symptomatology

We analyzed samples from our primary cohort of 177 previously SARS-CoV-2 infected HCW individuals in addition to the comparator cohorts comprising 54 HCs blood donors and 6 SLE patients. The median age of the primary cohort was 35 [IQR: 30-44] years, including 62 (35%) men with 38% of the total with a self-reported formal diagnosis of COVID-19 (**Supplemental Table 1**). Twenty-one COVID-19 related symptoms were scored based on their frequency of report by participants of the HCW survey. Overall, 13% (23/177) of the HCWs reported no symptoms, 36% (64/177) reported a mild overall symptom burden (i.e., having experienced up to 7 distinct symptom types), and 51% (90/177) reported a more than mild overall symptom burden (>7 symptoms). In the mild and the more than mild symptom burden groups, 33% and 51% reported PCR confirmed COVID-19 (**Supplemental Table 3**). The most frequently reported symptoms were fatigue (66%) and headache (62%), followed by muscle aches (57%), dry cough (56%) and nasal congestion (53%) (**Supplemental Table 3**).

### Diversity of IgG autoantibody reactivity

To target functional AABs that could present in response to SARS-CoV-2 infection, we used a high-throughput SeroTag^®^ AAB assay workflow. The degree of positive autoreactivity in all samples were defined as MFI value greater than 3-SD above the average of all pre-pandemic samples for each AAB readout (**Figure 1A**). A total of 160 out of 177 HCWs showed reactivity to at least one AABs. On average, 5.2 AABs were reactive in each HCW and only 2.3 AABs showed reactivity in HCs. Interestingly, 9% (n=17) HCWs did not show reactivity to any AABs, 36% (n=64) were positive for 1 to 2 AABs, 29% (n=52) were positive for 3 to 6 AABs and 25% (n=44) were positive for more than 6 AABs (**Supplement Table 4**). As expected, AABs were detected with much higher odds in SLE samples compared to HCs (P<0.0001, 95% 595.5 to 3319) and HCWs (P<0.0001, 95% -2843 to -119.8). The AABs detection rate was also higher in HCWs compare to HCs, although this difference did not meet the threshold of statistical significance (P=0.088, 95% -886 to 1837).

**Figure 1.**
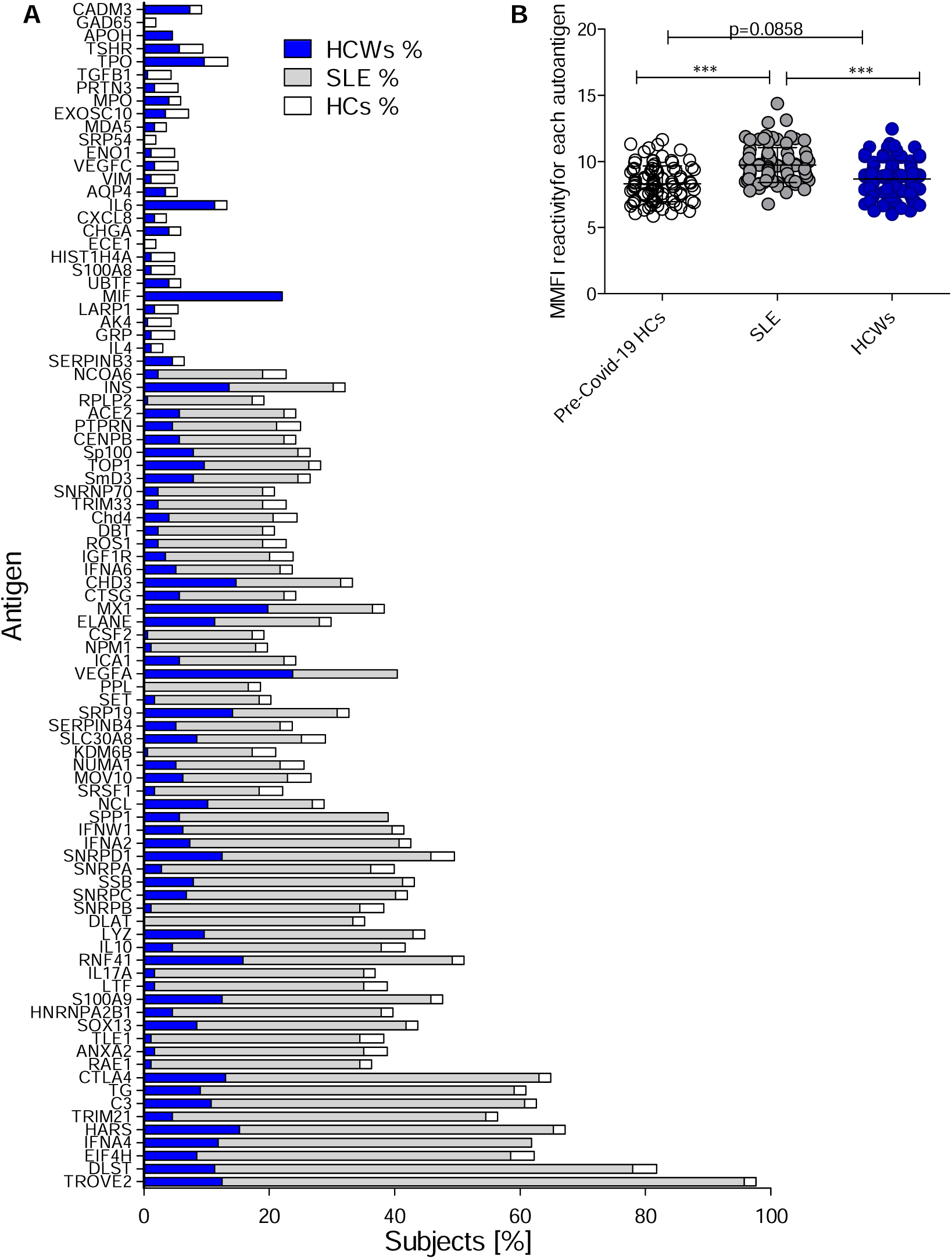
Autoantibody (AABs) expression among HCWs, pre-COVID-19 non-autoimmune controls and SLE, autoimmune positive group. **A**. Percentage of antigen reactive in study group. **B**. MFI values for each AABs in HCWs, HCs and SLE. Antigen abbreviation in supplement table 2.

We assessed whether autoantibody reactivity was associated with age, sex, or timing of the reported symptoms (**Supplement Table 4**). None of these variables were discernibly associated with IgG level. In analyses of the distinct AABs, examined individually, we found significant increase in IgG reactivity against 12 out of 91 antigens (including CHD3, CTLA4, HARS, IFNA4, INS, MIF, MX1, RNF41, S100A9, SRP19, TROVE2, VEGFA) in comparisons to HCs (**Supplement Table 5**). In addition, titers of anti-C3 (P=0.08), IL-6(P=0.07), ELANE (P=0.06), SMD3 (P=0.22), SP100 (P=0.22), SNRPD1 (P=0.12), and TOP1(P=0.12) were also elevated in HCW compare to HCs, but the elevation for each individual AAB did not meet the threshold of statistical significance. The majority of the AABs that showed statistical significance were previously described during Sars-CoV2 and other virus’ infection, including MX1, TOP1, SP100, SNRPD1, SNRPC, SSP1 (23-26). Thus, it is conceivable that SARS-CoV-2 infection may hijack host immune system in order to impair anti-viral immunity and further trigger a chronic inflammation characterized by anti-TROVE2, DLST, IFNA4, HARS, C3, CTLA4 and SMD3 found in our study. The identified antigens have been previously associated with the immune-mediated inflammatory diseases, like systemic lupus erythematosus (SLE)(27), dermatomyositis (DM)(28), multiple sclerosis (MS)(29), antisynthetase syndrome (ASS) (30) as well as predisposing factors known to exacerbate immune-related adverse events (irAEs) (31, 32). Remarkably, we also identified novel autoantibody targets implicated in organ-specific chronic inflammation, such as the thyroid in Graves’ disease (VEGFA, PTPRN, IGF1RTG) and the islet cell in type I diabetes (INS, S100A9, ICA1).

In Chi-square test for categorical variables of the primary HCW cohort, we examined pre-existing clinical and symptom traits in association with detected presence (AABs”+”) versus absence (ABBs”-”) of AABs defined based on the MIF parameter. The presence of AAB reactivity against CHD3 was associated with reported skin changes (AABs”-”(5%) *vs*. AABs”+” (19%) (P=0.035). Similarly, anti-SRP19 AABs was associated with shortness of breath symptom (AABs “-” (28%) vs AABs”+” (56%)(P=0.012), while anti-INS AABs was associated with reported fever (AABs “-” (48%) *vs*. AABs”+” (21%)(P=0.025)) and early occurring symptoms (AABs”-” (32%) *vs*. AABs”+” (67%)(P=0.006)); anti-IFNA4 AABs were increased in association with reported diarrhea (AABs”-” (27%) *vs*. AABs”+” (52%)(P=0.033)). IgG reactivity against CTLA4 was detected more frequently in (AABs”-” (49 (32%)) vs. AABs”+” (13(56%)(P=0.037) in males. There were no significant differences in the age, sex, race/ethnicity, and severity of the symptoms between HCWs who were positive or negative for the remaining AABs.

## DISCUSSION

Evidence has been rapidly accumulating, since the outbreak of COVID-19, relating SARS-CoV-2 infection with clinical manifestations that appear to directly emanate from viral tissue damage or indirectly induced by the antiviral immune response (33). A recent study by Gazzaruso *et. al*. has demonstrated the presence of autoimmune activation, based on detection of anti-nuclear autoantibodies (ANA) (36%) and lupus anti-coagulant (11%), in 45 cases of severe SARS-CoV-2 infection (34). Liu *et*.*al*. reported that antibodies against MDA5 was associated with amyopathic dermatomyositis following SARS-CoV-2 infection (18). Fujii *et. al*. reported a case-based review of two patients with severe respiratory failure due to COVID-19 who had high of anti-SSA/Ro antibody titer (35, 36). These studies suggest an autoimmune response in at least a subset of SARS-CoV-2 patients. Despite these data, it has remained still largely unknown how often AABs may be detected in SARS-CoV-2 recovering or recovered individuals without pre-existing immunologically mediated disorders. Hence, we tested whether SARS-CoV-2 infected individuals demonstrate the presence of distinct and specific autoantibody profiles that may be associated with classic autoimmune diseases or that may be manifestations of a naturally occurring stochastic repertoire. We used multiplex Luminex assay with 91 antigens and compared our cohort to SLE positive and healthy individuals who were not exposed to SARS-CoV2. According to our knowledge, we evaluated the largest number of protein antigens to characterize the prevalence and heterogeneity of the AABs signature in SARS-CoV-2 convalescent HCWs. We deliberately examined autoimmune reactivity to SARS-CoV-2 in the absence of extreme clinical disease to acknowledge the existence of AABs even among those who had mild-to-moderate or no symptoms during their illness, as a hallmark of ongoing long-COVID syndrome. The overall amount of detectable self-reactive IgG was >90% in HCWs, with on average 5.2 AABs positive in each HCW; this compared to 2.3 and 17.3 AABs positive in HCs and SLE blood donors, respectively. Importantly, detected biological autoimmunity in HCs is not always pathological, and can be observed in healthy people (37, 38). It has been suggested that the variety of plasma antibodies that could react to self and non-self-antigens regardless of external stimuli are essential components of self-defense in health and disease (39). Interestingly, 9% (n=17) HCWs did not show reactivity to any AABs, where 36% (n=64) were positive for 1 to2 AABs, 29% were positive for 3- to 6 AABs positive and 25% were positive for more than 6 AABs. It remains to be seen if any of the individuals we have investigated herein develop autoimmune diseases. Moreover, there was a striking paucity of some autoantibodies in HCWs not reactive in HCs, such as VEGFA, MIF, IFNA4, SPP1 and APOH.

MIF was previously described as a potential predictor for the outcome of critically ill patients and for acute respiratory distress syndrome (ARDS), a hallmark of severe COVID-19 disease (40). VEGF is identified as a principal proangiogenic factor that enhances the production of new blood vessels from existing vascular network. It has been reported before that viruses lack their own angiogenic factors; therefore, they exploit the cellular signaling machinery to upregulate the expression of VEGF and benefit from its physiological functions for their own pathogenesis. Understanding the interplay between viral infection and VEGF upregulation will pave the way to design targeted and effective therapeutic approaches for viral oncogenesis and severe diseases (41). In correlation to COVID-19, VEGFs concentrations were observed to be significantly higher in both ICU and non-ICU COVID-19 patients than in healthy controls (42). In agreement to another study, we also showed significant increased reactivity to interferon (specifically, IFNA4)(43, 44) as well as S100A9 (a circulating biomarker for inflammation-associated diseases, (45), in SARS-CoV-2-infected patients (46). S100A9 together with S100A8 are endogenous molecules released in response to environmental triggers and cellular damage. They are constitutively expressed in immune cells such as monocytes and neutrophils and their expression are upregulated under inflammatory conditions. (47). Impotently, as mentioned S100A8/S100A9 levels were reported to be associated with poor clinical outcomes such as significantly reduced survival time, especially in patients with severe pulmonary disease (48). SPP1 protein, also known as osteopenia, represents a multifunctional molecule playing a pivotal role in chronic inflammatory and autoimmune diseases(49). Its expression is increased in inflammatory bowel disease (IBD)(50). Similarly, autoimmune response to APOH, protein associated with autoimmune disorder called antiphospholipid syndrome(51), was also activated in HCWs in our data.

Our findings provide a strong rationale for a wider investigation of AABs in patients with COVID-19 as the data suggests humoral immunopathology is an intrinsic aspect of COVID-19 disease pathogenesis. We hypothesize that the group of seropositive but healthy individuals will develop autoimmune diseases, and the presence of AABs is prognostic marker that precedes clinical manifestation. On the other hand, some of AAB-positive, disease-free individuals may never develop clinical disease, and the presence of AABs may have no clinical significance for them. Although it is not known which of the above scenarios occurs, the current study provides important information. Long-term follow-up would be the best way to resolve this question.

## Data Availability

Requests for de-identified data may be directed to the corresponding authors (Jennifer E. Van Eyk, Susan Cheng, Justyna Fert-Bober) and will be reviewed by the Office of Research Administration at Cedars-Sinai Medical Center prior to issuance of data sharing agreements. Data limitations are designed to ensure patient and participant confidentiality.

## DATA AVAILABILITY

Requests for de-identified data may be directed to the corresponding authors (JEVE, SC, JFB) and will be reviewed by the Office of Research Administration at Cedars-Sinai Medical Center prior to issuance of data sharing agreements. Data limitations are designed to ensure patient and participant confidentiality.

## AUTHOR CONTRIBUTION

JFB, JEVE, JB, KS and SC conceived and designed the overall CORALE study. JFB, JB, KS, JEVE, and SC acquired the CORALE data. PB, JG AND H-D Z conceived the AABS analysis of the study, and YL, PB, JG, JFB, KS, JEVE, SC, and DM analyzed the data. JFB, SC, YL and DM drafted the manuscript, and all authors edited the manuscript.

## SUPPLEMENTAL MATERIAL

**Supplemental Table 1.** Demographic and clinical characteristics of HCWs.

|  | **Overall**<br>**(N=177)** | **Women**<br>**(N=115)** | **Men**<br>**(N=62)** | **P value** |
| --- | --- | --- | --- | --- |
| **Age (median [IQR])** | 35.00 [30.00, 44.00] | 35.00 [30.00, 46.50] | 34.50 [29.25, 40.75] | 0.53 |
| **Race/Ethnicity (%)** |  |  |  |  |
| Hispanic/Latinx | 50 (28.2) | 34 (29.6) | 16 (25.8) | 0.53 |
| Non-Hispanic Asian | 46 (26.0) | 30 (26.1) | 16 (25.8) |  |
| Non-Hispanic Black | 15 (8.5) | 11 (9.6) | 4 (6.5) |  |
| Non-Hispanic White | 56 (31.6) | 32 (27.8) | 24 (38.7) |  |
| Other | 10 (5.6) | 8 (7.0) | 2 (3.2) |  |
| **Medical Conditions** |  |  |  |  |
| Cancer | 2 (1.2) | 2 (1.8) | 0 (0.0) | 0.80 |
| Cardiovascular | 2 (1.2) | 1 (0.9) | 1 (1.7) | 1.00 |
| Chronic Obstructive Pulmonary Disease | 0 (0) | 0 (0) | 0 (0) | NA |
| Diabetes Mellitus | 5 (2.9) | 3 (2.7) | 2 (3.3) | 1.00 |
| Hypertension | 19 (11.0) | 13 (11.4) | 6 (10.2) | 1.00 |
| Immune | 3 (1.7) | 3 (2.7) | 0 (0.0) | 0.50 |
| Prior diagnosis of COVID-19 | 68 (38.4) | 42 (36.5) | 26 (41.9) | 0.59 |
| Smoking | 2 (1.1) | 1 (0.9) | 1 (1.6) | 1.00 |
| Vaping | 14 (7.9) | 6 (5.2) | 8 (12.9) | 0.13 |
| **Reported symptoms** |  |  |  |  |
| Chest pain | 36 (20.3) | 24 (20.9) | 12 (19.4) | 0.97 |
| Chills | 86 (48.6) | 52 (45.2) | 34 (54.8) | 0.29 |
| Conjunctivitis | 11 (6.2) | 5 (4.3) | 6 (9.7) | 0.28 |
| Dry cough | 100 (56.5) | 64 (55.7) | 36 (58.1) | 0.88 |
| Productive cough | 44 (24.9) | 32 (27.8) | 12 (19.4) | 0.29 |
| Diarrhea | 53 (29.9) | 31 (27.0) | 22 (35.5) | 0.31 |
| Fatigue | 117 (66.1) | 76 (66.1) | 41 (66.1) | 1.00 |
| Fever | 78 (44.1) | 47 (40.9) | 31 (50.0) | 0.31 |
| Headache | 110 (62.1) | 73 (63.5) | 37 (59.7) | 0.74 |
| Loss of appetite | 68 (38.4) | 49 (42.6) | 19 (30.6) | 0.16 |
| Muscle aches | 101 (57.1) | 66 (57.4) | 35 (56.5) | 1.00 |
| Nasal congestion | 93 (52.5) | 60 (52.2) | 33 (53.2) | 1.00 |
| Nausea | 47 (26.6) | 37 (32.2) | 10 (16.1) | 0.033 |
| Runny nose | 76 (42.9) | 55 (47.8) | 21 (33.9) | 0.10 |
| Shortness of breath | 57 (32.2) | 33 (28.7) | 24 (38.7) | 0.23 |
| Skin changes | 13 (7.3) | 9 (7.8) | 4 (6.5) | 0.97 |
| Smell and/or taste | 92 (52.0) | 61 (53.0) | 31 (50.0) | 0.82 |
| Sneezing | 83 (46.9) | 56 (48.7) | 27 (43.5) | 0.62 |
| Sore throat | 71 (40.1) | 48 (41.7) | 23 (37.1) | 0.66 |
| Stroke symptoms | 2 (1.1) | 1 (0.9) | 1 (1.6) | 1.00 |
| Vomiting | 16 (9.0) | 9 (7.8) | 7 (11.3) | 0.62 |

**Supplemental Table 2.**
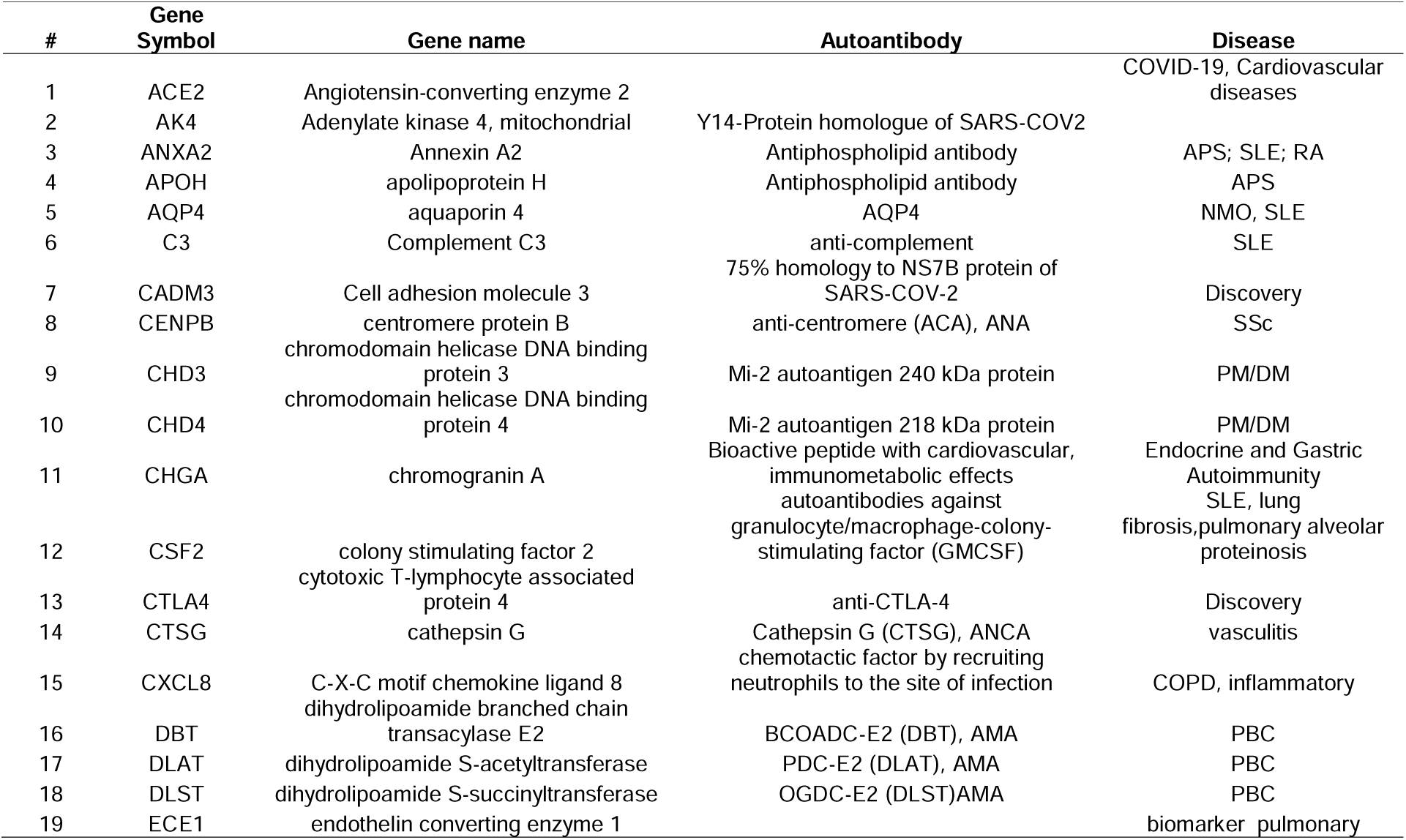
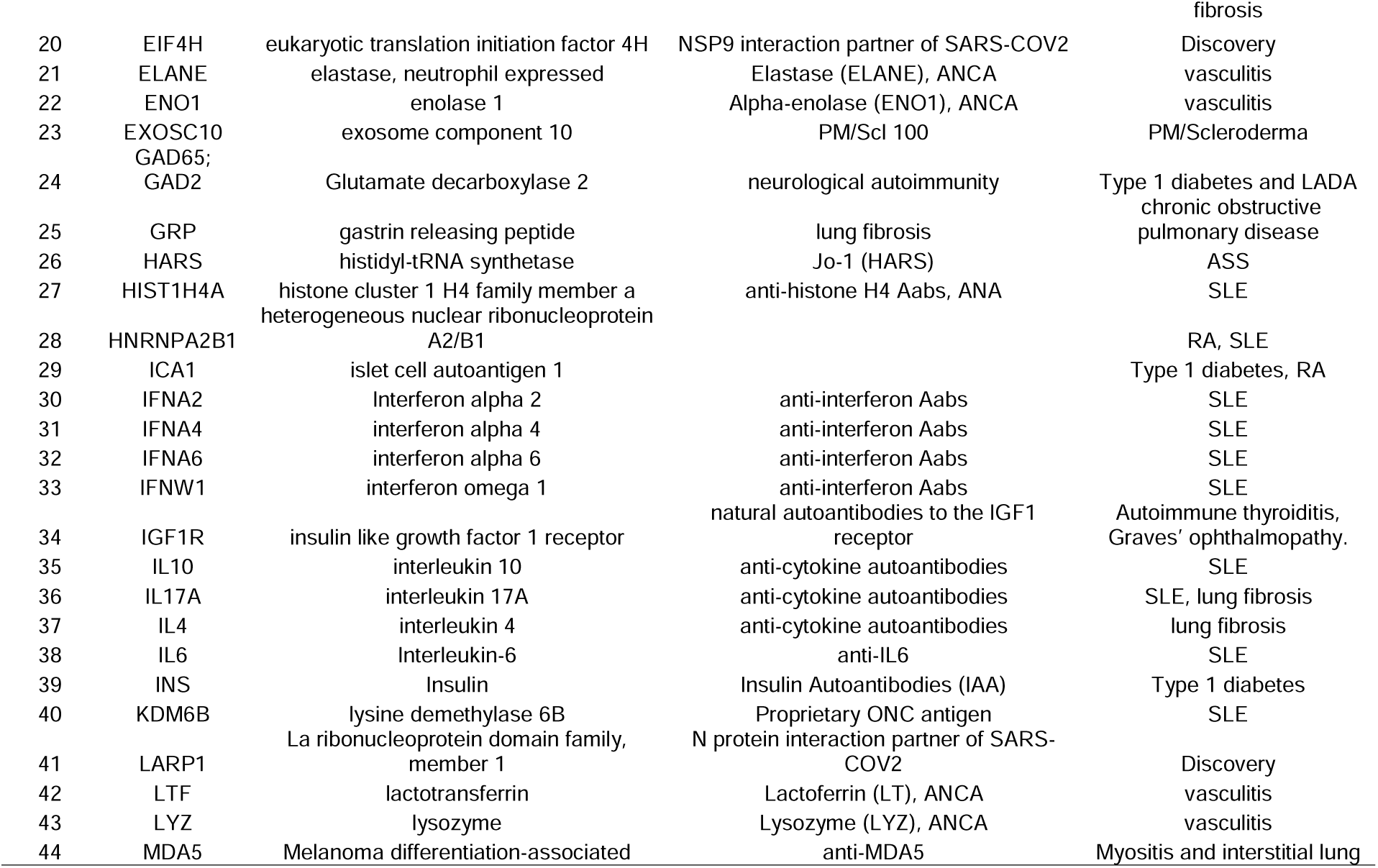
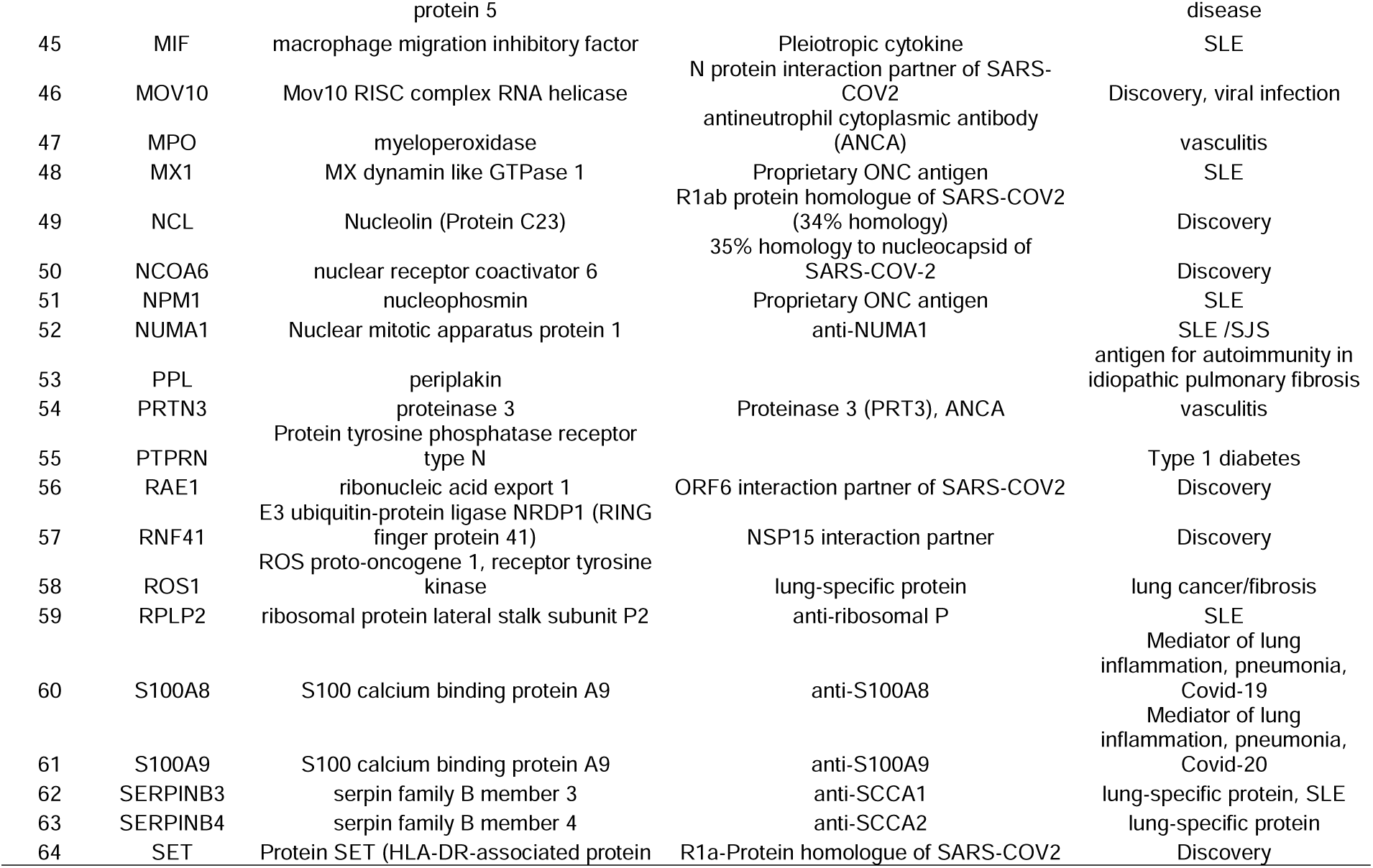
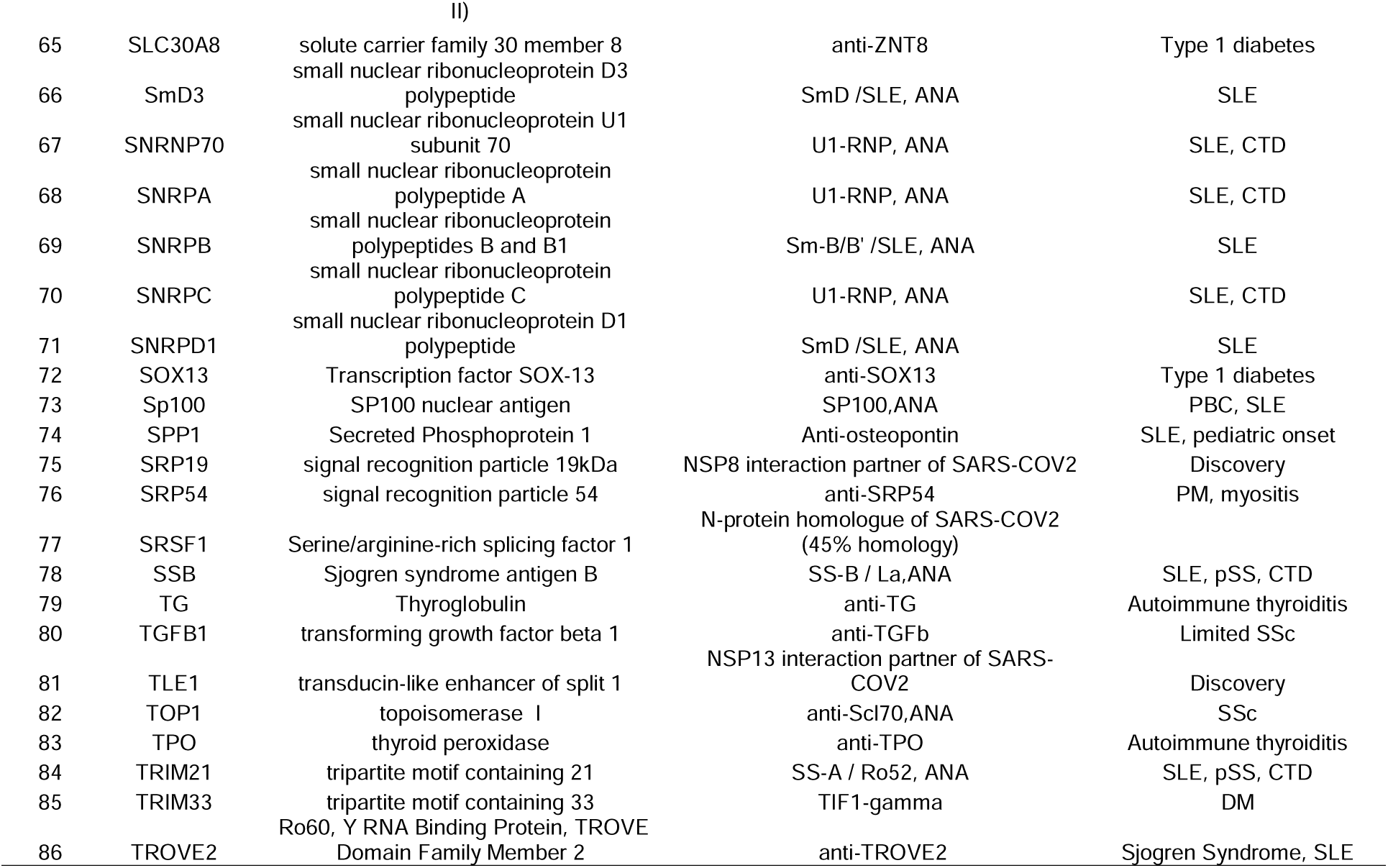
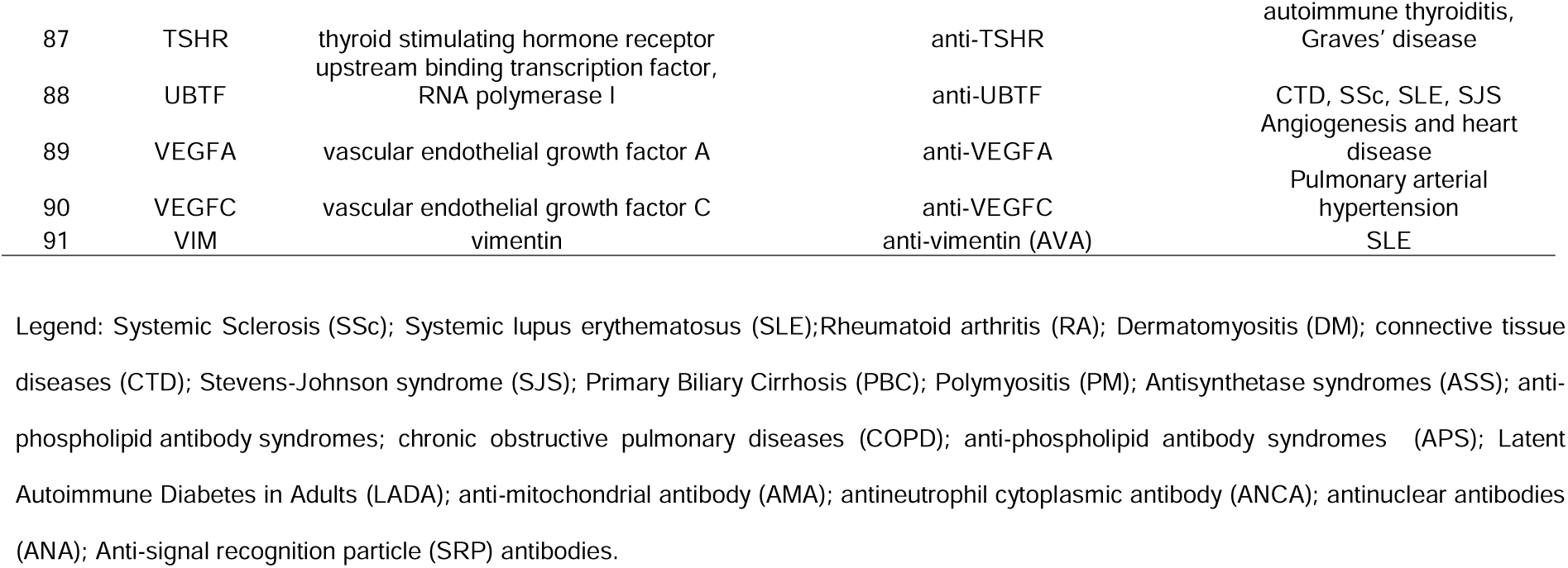
Composition of Antigen Array.

**Supplemental Table 3.** Symptoms scoring characteristics.

| **Category** | **WOMEN** | **MEN** | **PCR Confirmed** |
| --- | --- | --- | --- |
| **HCWs** | 115 | 62 | 68 (38.4%) |
| Asymptomatic | 15 (13%) | 8 (12.9%) | 1(4.3%) |
| Mild | 41 (35.7%) | 23 (37.1%) | 21 (32.8%) |
| More than mild | 59 (51.3%) | 31 (50%) | 46(51.1%) |
| **SLE** | 5 | 1 | NA |
| **Healthy Control** | 26 | 27 | NA |

**Supplemental Table 4.**
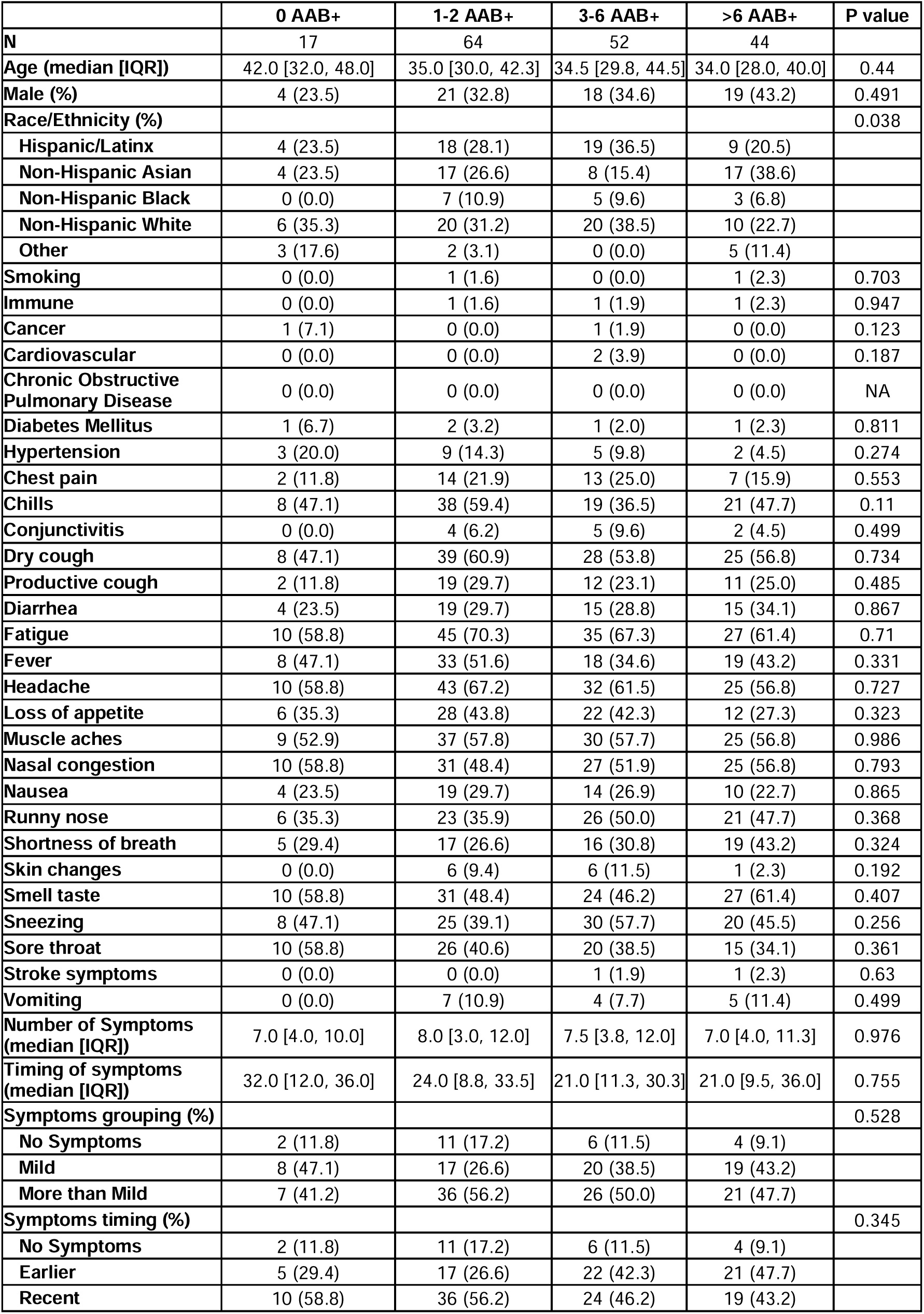
Comparison of characteristics with no ABB+ (N=17), 1-2 ABB+ (N=64), 3-6 ABB+ (N=52), and >6 ABB+ (N=44) in the HCWs.

|  | 0 AAB+ | 1-2 AAB+ | 3-6 AAB+ | >6 AAB+ | P value |
| --- | --- | --- | --- | --- | --- |
| **N** | 17 | 64 | 52 | 44 |  |
| **Age (median [IQR])** | 42.0 [32.0, 48.0] | 35.0 [30.0, 42.3] | 34.5 [29.8, 44.5] | 34.0 [28.0, 40.0] | 0.44 |
| **Male (%)** | 4 (23.5) | 21 (32.8) | 18 (34.6) | 19 (43.2) | 0.491 |
| **Race/Ethnicity (%)** |  |  |  |  | 0.038 |
| Hispanic/Latinx | 4 (23.5) | 18 (28.1) | 19 (36.5) | 9 (20.5) |  |
| Non-Hispanic Asian | 4 (23.5) | 17 (26.6) | 8 (15.4) | 17 (38.6) |  |
| Non-Hispanic Black | 0 (0.0) | 7 (10.9) | 5 (9.6) | 3 (6.8) |  |
| Non-Hispanic White | 6 (35.3) | 20 (31.2) | 20 (38.5) | 10 (22.7) |  |
| Other | 3 (17.6) | 2 (3.1) | 0 (0.0) | 5 (11.4) |  |
| **Smoking** | 0 (0.0) | 1 (1.6) | 0 (0.0) | 1 (2.3) | 0.703 |
| **Immune** | 0 (0.0) | 1 (1.6) | 1 (1.9) | 1 (2.3) | 0.947 |
| **Cancer** | 1 (7.1) | 0 (0.0) | 1 (1.9) | 0 (0.0) | 0.123 |
| **Cardiovascular** | 0 (0.0) | 0 (0.0) | 2 (3.9) | 0 (0.0) | 0.187 |
| **Chronic Obstructive Pulmonary Disease** | 0 (0.0) | 0 (0.0) | 0 (0.0) | 0 (0.0) | NA |
| **Diabetes Mellitus** | 1 (6.7) | 2 (3.2) | 1 (2.0) | 1 (2.3) | 0.811 |
| **Hypertension** | 3 (20.0) | 9 (14.3) | 5 (9.8) | 2 (4.5) | 0.274 |
| **Chest pain** | 2 (11.8) | 14 (21.9) | 13 (25.0) | 7 (15.9) | 0.553 |
| **Chills** | 8 (47.1) | 38 (59.4) | 19 (36.5) | 21 (47.7) | 0.11 |
| **Conjunctivitis** | 0 (0.0) | 4 (6.2) | 5 (9.6) | 2 (4.5) | 0.499 |
| **Dry cough** | 8 (47.1) | 39 (60.9) | 28 (53.8) | 25 (56.8) | 0.734 |
| **Productive cough** | 2 (11.8) | 19 (29.7) | 12 (23.1) | 11 (25.0) | 0.485 |
| **Diarrhea** | 4 (23.5) | 19 (29.7) | 15 (28.8) | 15 (34.1) | 0.867 |
| **Fatigue** | 10 (58.8) | 45 (70.3) | 35 (67.3) | 27 (61.4) | 0.71 |
| **Fever** | 8 (47.1) | 33 (51.6) | 18 (34.6) | 19 (43.2) | 0.331 |
| **Headache** | 10 (58.8) | 43 (67.2) | 32 (61.5) | 25 (56.8) | 0.727 |
| **Loss of appetite** | 6 (35.3) | 28 (43.8) | 22 (42.3) | 12 (27.3) | 0.323 |
| **Muscle aches** | 9 (52.9) | 37 (57.8) | 30 (57.7) | 25 (56.8) | 0.986 |
| **Nasal congestion** | 10 (58.8) | 31 (48.4) | 27 (51.9) | 25 (56.8) | 0.793 |
| **Nausea** | 4 (23.5) | 19 (29.7) | 14 (26.9) | 10 (22.7) | 0.865 |
| **Runny nose** | 6 (35.3) | 23 (35.9) | 26 (50.0) | 21 (47.7) | 0.368 |
| **Shortness of breath** | 5 (29.4) | 17 (26.6) | 16 (30.8) | 19 (43.2) | 0.324 |
| **Skin changes** | 0 (0.0) | 6 (9.4) | 6 (11.5) | 1 (2.3) | 0.192 |
| **Smell taste** | 10 (58.8) | 31 (48.4) | 24 (46.2) | 27 (61.4) | 0.407 |
| **Sneezing** | 8 (47.1) | 25 (39.1) | 30 (57.7) | 20 (45.5) | 0.256 |
| **Sore throat** | 10 (58.8) | 26 (40.6) | 20 (38.5) | 15 (34.1) | 0.361 |
| **Stroke symptoms** | 0 (0.0) | 0 (0.0) | 1 (1.9) | 1 (2.3) | 0.63 |
| **Vomiting** | 0 (0.0) | 7 (10.9) | 4 (7.7) | 5 (11.4) | 0.499 |
| **Number of Symptoms (median [IQR])** | 7.0 [4.0, 10.0] | 8.0 [3.0, 12.0] | 7.5 [3.8, 12.0] | 7.0 [4.0, 11.3] | 0.976 |
| **Timing of symptoms (median [IQR])** | 32.0 [12.0, 36.0] | 24.0 [8.8, 33.5] | 21.0 [11.3, 30.3] | 21.0 [9.5, 36.0] | 0.755 |
| **Symptoms grouping (%)** |  |  |  |  | 0.528 |
| No Symptoms | 2 (11.8) | 11 (17.2) | 6 (11.5) | 4 (9.1) |  |
| Mild | 8 (47.1) | 17 (26.6) | 20 (38.5) | 19 (43.2) |  |
| More than Mild | 7 (41.2) | 36 (56.2) | 26 (50.0) | 21 (47.7) |  |
| **Symptoms timing (%)** |  |  |  |  | 0.345 |
| No Symptoms | 2 (11.8) | 11 (17.2) | 6 (11.5) | 4 (9.1) |  |
| Earlier | 5 (29.4) | 17 (26.6) | 22 (42.3) | 21 (47.7) |  |
| Recent | 10 (58.8) | 36 (56.2) | 24 (46.2) | 19 (43.2) |  |

**Supplemental Table 5.** Chi-square analysis of correlation of AABs in HCWs compared to HCs (non-SLE pre-pandemic samples).

| **Autoantibody** | **Control (N=53)** | **HCWs (N=177)** | **P value** |
| --- | --- | --- | --- |
| ACE2 | 1 (1.9) | 10 (5.6) | 0.45 |
| AK4 | 2 (3.8) | 1 (0.6) | 0.26 |
| ANXA2 | 2 (3.8) | 3 (1.7) | 0.71 |
| APOH | 1 (1.9) | 8 (4.5) | 0.64 |
| AQP4 | 1 (1.9) | 6 (3.4) | 0.92 |
| C3 | 1 (1.9) | 19 (10.7) | 0.08 |
| CADM3 | 1 (1.9) | 13 (7.3) | 0.26 |
| CENPB | 1 (1.9) | 10 (5.6) | 0.45 |
| CHD3 | 1 (1.9) | 26 (14.7) | 0.022 |
| CHD4 | 2 (3.8) | 7 (4.0) | 1.00 |
| CHGA | 1 (1.9) | 7 (4.0) | 0.77 |
| CSF2 | 1 (1.9) | 1 (0.6) | 0.95 |
| CTLA4 | 1 (1.9) | 23 (13.0) | 0.039 |
| CTSG | 1 (1.9) | 10 (5.6) | 0.45 |
| CXCL8 | 1 (1.9) | 3 (1.7) | 1.00 |
| DBT | 1 (1.9) | 4 (2.3) | 1.00 |
| DLAT | 1 (1.9) | 0 (0.0) | 0.52 |
| DLST | 2 (3.8) | 20 (11.3) | 0.17 |
| ECE1 | 1 (1.9) | 0 (0.0) | 0.52 |
| EIF4H | 2 (3.8) | 15 (8.5) | 0.40 |
| ELANE | 1 (1.9) | 20 (11.3) | 0.07 |
| ENO1 | 2 (3.8) | 2 (1.1) | 0.49 |
| EXOSC10 | 2 (3.8) | 6 (3.4) | 1.00 |
| GAD65 | 1 (1.9) | 0 (0.0) | 0.52 |
| GRP | 2 (3.8) | 2 (1.1) | 0.49 |
| HARS | 1 (1.9) | 27 (15.3) | 0.018 |
| HIST1H4A | 2 (3.8) | 2 (1.1) | 0.49 |
| HNRNPA2B1 | 1 (1.9) | 8 (4.5) | 0.64 |
| ICA1 | 1 (1.9) | 10 (5.6) | 0.45 |
| IFNA2 | 1 (1.9) | 13 (7.3) | 0.26 |
| IFNA4 | 0 (0.0) | 21 (11.9) | 0.018 |
| IFNA6 | 1 (1.9) | 9 (5.1) | 0.54 |
| IFNW1 | 1 (1.9) | 11 (6.2) | 0.37 |
| IGF1R | 2 (3.8) | 6 (3.4) | 1.00 |
| IL10 | 2 (3.8) | 8 (4.5) | 1.00 |
| IL17A | 1 (1.9) | 3 (1.7) | 1.00 |
| IL4 | 1 (1.9) | 2 (1.1) | 1.00 |
| IL6 | 1 (1.9) | 20 (11.3) | 0.07 |
| INS | 1 (1.9) | 24 (13.6) | 0.032 |
| KDM6B | 2 (3.8) | 1 (0.6) | 0.26 |
| LARP1 | 2 (3.8) | 3 (1.7) | 0.71 |
| LTF | 2 (3.8) | 3 (1.7) | 0.71 |
| LYZ | 1 (1.9) | 17 (9.6) | 0.12 |
| MDA5 | 1 (1.9) | 3 (1.7) | 1.00 |
| MIF | 0 (0.0) | 39 (22.0) | <0.001 |
| MOV10 | 2 (3.8) | 11 (6.2) | 0.74 |
| MPO | 1 (1.9) | 7 (4.0) | 0.77 |
| MX1 | 1 (1.9) | 35 (19.8) | 0.003 |
| NCL | 1 (1.9) | 18 (10.2) | 0.10 |
| NCOA6 | 2 (3.8) | 4 (2.3) | 0.91 |
| NPM1 | 1 (1.9) | 2 (1.1) | 1.00 |
| NUMA1 | 2 (3.8) | 9 (5.1) | 0.98 |
| PPL | 1 (1.9) | 0 (0.0) | 0.52 |
| PRTN3 | 2 (3.8) | 3 (1.7) | 0.71 |
| PTPRN | 2 (3.8) | 8 (4.5) | 1.00 |
| RAE1 | 1 (1.9) | 2 (1.1) | 1.00 |
| RNF41 | 1 (1.9) | 28 (15.8) | 0.014 |
| ROS1 | 2 (3.8) | 4 (2.3) | 0.91 |
| RPLP2 | 1 (1.9) | 1 (0.6) | 0.95 |
| S100A8 | 2 (3.8) | 2 (1.1) | 0.49 |
| S100A9 | 1 (1.9) | 22 (12.4) | 0.047 |
| SERPINB3 | 1 (1.9) | 8 (4.5) | 0.64 |
| SERPINB4 | 1 (1.9) | 9 (5.1) | 0.54 |
| SET | 1 (1.9) | 3 (1.7) | 1.00 |
| SLC30A8 | 2 (3.8) | 15 (8.5) | 0.40 |
| SMD3 | 1 (1.9) | 14 (7.9) | 0.22 |
| SNRNP70 | 1 (1.9) | 4 (2.3) | 1.00 |
| SNRPA | 2 (3.8) | 5 (2.8) | 1.00 |
| SNRPB | 2 (3.8) | 2 (1.1) | 0.49 |
| SNRPC | 1 (1.9) | 12 (6.8) | 0.31 |
| SNRPD1 | 2 (3.8) | 22 (12.4) | 0.12 |
| SOX13 | 1 (1.9) | 15 (8.5) | 0.18 |
| SP100 | 1 (1.9) | 14 (7.9) | 0.22 |
| SPP1 | 0 (0.0) | 10 (5.6) | 0.17 |
| SRP19 | 1 (1.9) | 25 (14.1) | 0.026 |
| SRP54 | 1 (1.9) | 0 (0.0) | 0.52 |
| SRSF1 | 2 (3.8) | 3 (1.7) | 0.71 |
| SSB | 1 (1.9) | 14 (7.9) | 0.22 |
| TG | 1 (1.9) | 16 (9.0) | 0.15 |
| TGFB1 | 2 (3.8) | 1 (0.6) | 0.26 |
| TLE1 | 2 (3.8) | 2 (1.1) | 0.49 |
| TOP1 | 1 (1.9) | 17 (9.6) | 0.12 |
| TPO | 2 (3.8) | 17 (9.6) | 0.29 |
| TRIM21 | 1 (1.9) | 8 (4.5) | 0.64 |
| TRIM33 | 2 (3.8) | 4 (2.3) | 0.91 |
| TROVE2 | 1 (1.9) | 22 (12.4) | 0.047 |
| TSHR | 2 (3.8) | 10 (5.6) | 0.85 |
| UBTF | 1 (1.9) | 7 (4.0) | 0.77 |
| VEGFA | 0 (0.0) | 42 (23.7) | <0.001 |
| VEGFC | 2 (3.8) | 3 (1.7) | 0.71 |
| VIM | 2 (3.8) | 2 (1.1) | 0.49 |

## REFERENCES

1. Lerma LA, et al. (2020) Prevalence of autoantibody responses in acute coronavirus disease 2019 (COVID-19). Journal of translational autoimmunity 3:100073.

2. Ingraham NE, et al. (2020) Immunomodulation in COVID-19. The Lancet Respiratory Medicine 8(6):544–546.

3. Tenforde MW, et al. (2020) Symptom Duration and Risk Factors for Delayed Return to Usual Health Among Outpatients with COVID-19 in a Multistate Health Care Systems Network - United States, March-June 2020. MMWR Morb Mortal Wkly Rep 69(30):993–998.

4. Ahmad T, et al. (2020) COVID-19: The Emerging Immunopathological Determinants for Recovery or Death. Frontiers in Microbiology 11(2815).

5. Mantovani Cardoso E, Hundal J, Feterman D, & Magaldi J (2020) Concomitant new diagnosis of systemic lupus erythematosus and COVID-19 with possible antiphospholipid syndrome. Just a coincidence? A case report and review of intertwining pathophysiology. Clin Rheumatol 39(9):2811–2815.

6. Toscano G, et al. (2020) Guillain-Barré Syndrome Associated with SARS-CoV-2. The New England journal of medicine 382(26):2574–2576.

7. Zhao H, Shen D, Zhou H, Liu J, & Chen S (2020) Guillain-Barré syndrome associated with SARS-CoV-2 infection: causality or coincidence? The Lancet. Neurology 19(5):383–384.

8. Becker RC (2020) COVID-19-associated vasculitis and vasculopathy. Journal of thrombosis and thrombolysis 50(3):499–511.

9. Palao M, et al. (2020) Multiple sclerosis following SARS-CoV-2 infection. Multiple sclerosis and related disorders 45:102377.

10. Bastard P, et al. (2020) Autoantibodies against type I IFNs in patients with life-threatening COVID-19. Science (New York, N.Y.) 370(6515).

11. Ebinger JE, et al. (2021) Seroprevalence of antibodies to SARS-CoV-2 in healthcare workers: a cross-sectional study. BMJ open 11(2):e043584.

12. Ebinger JE, et al. (2020) An Opportune and Relevant Design for Studying the Health Trajectories of Healthcare Workers. medRxiv:2020.2006.2030.20140046.

13. Bryan A, et al. (2020) Performance Characteristics of the Abbott Architect SARS-CoV-2 IgG Assay and Seroprevalence in Boise, Idaho. Journal of clinical microbiology 58(8).

14. Harris PA, et al. (2019) The REDCap consortium: Building an international community of software platform partners. Journal of biomedical informatics 95:103208.

15. Budde P, et al. (2016) Multiparametric detection of autoantibodies in systemic lupus erythematosus. Lupus 25(8):812–822.

16. Corman VM, et al. (2020) Detection of 2019 novel coronavirus (2019-nCoV) by real-time RT-PCR. Euro Surveill 25(3):2000045.

17. Gordon DE, et al. (2020) A SARS-CoV-2-Human Protein-Protein Interaction Map Reveals Drug Targets and Potential Drug-Repurposing. bioRxiv : the preprint server for biology:2020.2003.2022.002386.

18. Liu C, et al. (2020) Augmentation of anti-MDA5 antibody implies severe disease in COVID-19 patients. medRxiv:2020.2007.2029.20164780.

19. Hassan SS, et al. (2021) Notable sequence homology of the ORF10 protein introspects the architecture of SARS-CoV-2. Int J Biol Macromol 181:801–809.

20. Macdonald IK, et al. (2012) Development and validation of a high throughput system for discovery of antigens for autoantibody detection. PloS one 7(7):e40759.

21. Carpenter B, et al. (2017) Stan: A Probabilistic Programming Language. 2017 76(1):32.

22. Dong Q & Gao X (2020) Bayesian estimation of the seroprevalence of antibodies to SARS-CoV-2. JAMIA open 3(4):496–499.

23. Collados Rodríguez M (2021) The Fate of Speckled Protein 100 (Sp100) During Herpesviruses Infection. Frontiers in Cellular and Infection Microbiology 10(900).

24. Laise P, et al. (2020) The Host Cell ViroCheckpoint: Identification and Pharmacologic Targeting of Novel Mechanistic Determinants of Coronavirus-Mediated Hijacked Cell States. bioRxiv : the preprint server for biology:2020.2005.2012.091256.

25. Takahashi K, et al. (2013) DNA topoisomerase 1 facilitates the transcription and replication of the Ebola virus genome. Journal of virology 87(16):8862–8869.

26. Bost P, et al. (2020) Deciphering the state of immune silence in fatal COVID-19 patients. medRxiv:2020.2008.2010.20170894.

27. Mahler M, Stinton LM, & Fritzler MJ (2005) Improved Serological Differentiation between Systemic Lupus Erythematosus and Mixed Connective Tissue Disease by Use of an SmD3 Peptide-Based Immunoassay. Clinical and Vaccine Immunology 12(1):107–113.

28. López de Padilla CM & Niewold TB (2016) The type I interferons: Basic concepts and clinical relevance in immune-mediated inflammatory diseases. Gene 576(1 Pt 1):14–21.

29. Ingram G, et al. (2014) Complement activation in multiple sclerosis plaques: an immunohistochemical analysis. Acta Neuropathologica Communications 2(1):53.

30. Adams RA, et al. (2021) Serum-circulating His-tRNA synthetase inhibits organ-targeted immune responses. Cellular & Molecular Immunology 18(6):1463–1475.

31. Kong Y-CM & Flynn JC (2014) Opportunistic Autoimmune Disorders Potentiated by Immune-Checkpoint Inhibitors Anti-CTLA-4 and Anti-PD-1. Frontiers in immunology 5:206–206.

32. Wang JY, Zhang W, Roehrl MW, Roehrl VB, & Roehrl MH (2021) An Autoantigen Atlas from Human Lung HFL1 Cells Offers Clues to Neurological and Diverse Autoimmune Manifestations of COVID-19. bioRxiv : the preprint server for biology:2021.2001.2024.427965.

33. Talotta R & Robertson E (2020) Autoimmunity as the comet tail of COVID-19 pandemic. World J Clin Cases 8(17):3621–3644.

34. Gazzaruso C, et al. (2020) High prevalence of antinuclear antibodies and lupus anticoagulant in patients hospitalized for SARS-CoV2 pneumonia. Clin Rheumatol 39(7):2095–2097.

35. Fujii H, et al. (2020) High levels of anti-SSA/Ro antibodies in COVID-19 patients with severe respiratory failure: a case-based review : High levels of anti-SSA/Ro antibodies in COVID-19. Clin Rheumatol 39(11):3171–3175.

36. Halpert G & Shoenfeld Y (2020) SARS-CoV-2, the autoimmune virus. Autoimmunity reviews 19(12):102695–102695.

37. Palma J, Tokarz-Deptuła B, Deptuła J, & Deptuła W (2018) Natural antibodies - facts known and unknown. Cent Eur J Immunol 43(4):466–475.

38. Haller-Kikkatalo K, et al. (2017) Demographic associations for autoantibodies in disease-free individuals of a European population. Scientific Reports 7(1):44846.

39. Neiman M, et al. (2019) Individual and stable autoantibody repertoires in healthy individuals. Autoimmunity 52(1):1–11.

40. Bleilevens C, et al. (2021) Macrophage Migration Inhibitory Factor (MIF) Plasma Concentration in Critically Ill COVID-19 Patients: A Prospective Observational Study. Diagnostics 11(2).

41. Alkharsah KR (2018) VEGF Upregulation in Viral Infections and Its Possible Therapeutic Implications. International journal of molecular sciences 19(6).

42. Yin XX, Zheng XR, Peng W, Wu ML, & Mao XY (2020) Vascular Endothelial Growth Factor (VEGF) as a Vital Target for Brain Inflammation during the COVID-19 Outbreak. ACS chemical neuroscience 11(12):1704–1705.

43. Xiao C-X, et al. (2015) Exome sequencing identifies novel compound heterozygous IFNA4 and IFNA10 mutations as a cause of impaired function in Crohn’s disease patients. Scientific Reports 5(1):10514.

44. Mudter J, et al. (2008) The transcription factor IFN regulatory factor-4 controls experimental colitis in mice via T cell-derived IL-6. The Journal of clinical investigation 118(7):2415–2426.

45. Wang S, et al. (2018) S100A8/A9 in Inflammation. Frontiers in Immunology 9(1298).

46. Guo Q, et al. (2021) Induction of alarmin S100A8/A9 mediates activation of aberrant neutrophils in the pathogenesis of COVID-19. Cell Host & Microbe 29(2):222–235.e224.

47. Cher JZB, et al. (2018) Alarmins in Frozen Shoulder: A Molecular Association Between Inflammation and Pain. The American journal of sports medicine 46(3):671–678.

48. Mahler M, Meroni PL, Infantino M, Buhler KA, & Fritzler MJ (2021) Circulating Calprotectin as a Biomarker of COVID-19 Severity. Expert review of clinical immunology 17(5):431–443.

49. Schlecht A, et al. (2021) Secreted Phosphoprotein 1 Expression in Retinal Mononuclear Phagocytes Links Murine to Human Choroidal Neovascularization. Frontiers in Cell and Developmental Biology 8(1641).

50. Glas J, et al. (2011) The Role of Osteopontin (OPN/SPP1) Haplotypes in the Susceptibility to Crohn’s Disease. PloS one 6(12):e29309.

51. Tan Y, Bian Y, Song Y, Zhang Q, & Wan X (2021) Exosome-Contained APOH Associated With Antiphospholipid Syndrome. Frontiers in Immunology 12(1237).

